# Associations between parental attitudes towards mental illness and self-reported mental health among young people: Evidence from the Health Survey for England

**DOI:** 10.1101/2022.11.21.22282163

**Authors:** Thierry Gagné, Claire Henderson, Anne McMunn

## Abstract

**Background:** The prevalence of mental health problems among young adults has rapidly increased over the past decade. The argument that reductions in stigma lead to less under-reporting over time is often presented as a potential explanation. As a first step towards understanding how stigma influence self-reporting in this age group, we examine the extent to which parents’ attitudes are related to young people’s self-reported mental health.

**Methods:** We leveraged the household design of the 2014 Health Survey for England to test whether mothers’ (complete-case *n* = 630) and fathers’ (*n* = 428) prejudice and tolerance towards people with mental illness is associated with the self-reporting of any specific mental disorder and non-specific psychological distress (GHQ-12) in participants aged 13-24. Associations were tested in random-intercept Poisson models (nesting participants in households) adjusting for parents’ sociodemographics and mental health, and participants’ own sociodemographics.

**Results:** Mothers were on average less prejudiced (81.2 versus 74.1 out of 100) but as tolerant (72.0 versus 70.0 out of 100) as fathers. In fully-adjusted models: 1) those with a less prejudiced (PR for a one-unit increase = 1.036, 95%CI 1.007-1.066) and more tolerant (PR = 1.038, 95%CI 1.011-1.066) mother had a higher probability of reporting a mental disorder; 2) those with a less prejudiced (PR = 1.034, 95%CI 1.006-1.062) father had a higher probability of reporting a mental disorder; 3) those with a more tolerant father also had a higher probability of reporting a high level of psychological distress (PR = 1.024, 95%CI 1.008-1.041).

**Conclusion:** Parents’ attitudes were associated with their children’s mental health, more so with specific mental disorders compared with non-specific psychological distress. New data collection efforts are needed to understand changes in parental attitudes over time and its relationship with self-reporting among young people.

## 1. INTRODUCTION

A large evidence base across Canada, the United States, and the United Kingdom has highlighted a worrisome increase in mental health problems among young people over the past decade, disproportionately affecting women and those from more socioeconomically disadvantaged backgrounds. Studies focussed on changes since the start of the COVID-19 pandemic also found that young people reported a higher increase in psychological distress across age groups, with unequal experiences noted again in more disadvantaged groups.^1,2^

A common challenge to the notion that population levels of mental health have been worsening over time is “de-stigmatisation”, i.e.: 1) the presence of public stigma towards mental illness leads to the under-reporting of mental health problems; 2) improvements in public stigma over time should lead to decreasing levels of under-reporting and the “artificial” inflation of the prevalence of mental health problems.^3^ Supporting this, a number of studies using different techniques (i.e., analysis of repeated attitudinal surveys and document analysis of traditional media outputs) found improvements in stigma-related indicators across Canada, the U.S., and the UK over the same period that prevalence estimates have worsened.^4–9^

The extent to which time trends in public stigma and mental health are related, however, remains unclear, with at least one study finding no overlap between changes in stigma-related indicators and self-reported mental disorders across English regions over the past decade.^10^ Part of the problem in disentangling this mechanism concerns the different levels at which stigma operates, and the extent to which each of these may affect self-reporting.^11^ A number of studies examined the variability of stigma-related indicators and mental health problems in England and found that while the local and regional levels may not have a substantive role, the household level may influence the relationship between stigma and self-reporting to a meaningful extent.^12–15^

A large body of work has explored the role of parents’ conditions and practices in their children’s mental healht and related service use.^16,17^ Research on the specific contribution of parents’ stigmatising attitudes has been relatively small and focussed on children’s help-seeking practices.^18^ One U.S. study found that boys aged 13-14 were less willing to seek mental health services if their parents disapproved of such services compared with girls.^19^ Other U.S. studies found that this relationship varied by ethnicity, and was stronger among Hispanic youth.^20,21^ In England, one study found that young people aged 9-18 were more likely to have used mental health care services in the past year if their caregivers had less stigmatising attitudes, especially if they had not used such services themselves.^22^ A qualitative study on adolescent self-harm also found that parents’ support was likely to be associated with the willingness of young people to disclose their behaviour and seek help.^23^ One issue raised concerned the timing of this influence, i.e., whether parents’ attitudes continued having a direct effect after the end of adolescence. For instance, Yap and colleagues examined the correlates of help-seeking intentions and beliefs about the usefulness of health care services among Australian youth (ages 12-17) and young adults (ages 18-25), and found that parents’ attitudes were not associated with these variables in multivariable models that included participants’ own attitudes.^24^

### 1.1. Objectives

Few studies have explored the extent to which parents’ attitudes directly relate to young people’s self-reporting of mental health problems. If parents become less prejudiced over time, their children should become more likely to report a problem when there is one. To shed light on this issue, this study examines the extent to which parents’ attitudes are associated with their children’s self-reported mental health. Specifically, we build on the household design and mental health focus of a survey representative of the population in England, the 2014 Health Survey for England (HSE), to test the role of parents’ attitudes towards people with mental illness among young people aged 13-24 living at home. Importantly, we consider two outcomes to explore whether parental attitudes have a different magnitude of association when the outcome is a self-disclosed specific condition or a threshold based on a multi-item scale (which is the most common measurement approach across epidemiological studies).^25^ In keeping with potential gender differences in the role of parents’ attitudes, we distinguish between the attitudes of fathers and mothers.^26,27^

## 2. METHODS

### 2.1. Data

The HSE comprises a series of annual surveys started in 1991 and is designed to monitor trends in the nation’s health.^28^ The HSE is currently commissioned by NHS Digital and carried out by the Joint Health Surveys Unit of NatCen Social Research and the Department of Epidemiology and Public Health at University College London. The HSE uses a complex stratified, clustered design to ensure sufficient numbers of participants across regions and estimates representative of the English population. HSE annual waves include a core questionnaire and often implement an additional focus by modifying its sample design and/or core content, with 2014 including a one-time module on mental health.^29^

The achieved sample for the 2014 survey was 2,003 children aged 0-15 and 8,077 adults aged 16 and over, with an estimated individual response rate of 55%.^30^ This included 1,116 participants aged 13-24. The lower age bracket was chosen based on the availability of mental health measures across age groups. From this sample, we were able to match 861 participants (as natural, adopted, foster, step-son/daughter, or son/daughter-in law) from 668 households with at least one parent. This led to 800 participants linked to a mother (97% natural) and 557 participants linked to a father (88% natural). We randomly selected one parent to be the mother/father for participants in the few households where two parents were identified as mothers or fathers.

### 2.2. Measures

We used two self-reported outcomes to capture mental health: 1) presence of any specific mental disorder, measured during the main interview, and 2) presence of signficant psychological distress, measured in the subsequent self-completion questionnaire.

#### Outcome 1: Presence of any mental disorder

The presence of self-reported mental disorder(s) (SRMDs) (Yes / No) was derived by the HSE data management team based on open responses to a follow-up question with regard to longstanding health conditions (*“Do you have any physical or mental health conditions or illnesses lasting or expected to last 12 months or more?”*). The coding frame used by the HSE for this variable includes both mental illnesses (e.g., alcoholism, drug addiction, anxiety, depression, schizophrenia) and mental handicaps (see conditions in Supplementary Table 1).

#### Outcome 2: Psychological distress

Psychological distress (Yes / No) was measured using the 12-item General Health Questionnaire (GHQ-12), a validated screening tool for identifying non-psychotic and minor psychiatric disorders in the general population. Each item uses a 4-point Likert-type scale (0-3) to assess the severity of a problem over the past few weeks (e.g., *“Have you recently been feeling reasonably happy, all things considered?”*; see items in Supplementary Table 2). In keeping with thresholds commonly used to define “caseness”, participants were defined to display a significant level of distress if they selected one of the two bottom response options on 4 or more out of the 12 items (yes or no).^31^

#### Exposure: Parents’ attitudes towards mental illness

The 12-item Community Attitudes towards Mental Illness (CAMI-12) scale is a shortened version of a longstanding scale initially developed to capture neighbourhood opposition to community-based mental health facilities in the 1970s.^32^ Psychometric analysis of the CAMI-12 scale supports two different scores representing prejudice (e.g., *“People with mental illness don’t deserve our sympathy”*) and tolerance (e.g., *“Virtually anyone can become mentally ill”*) (see items and reliability coefficients in Supplementary Table 3). CAMI items’ response scale vary from 1 – Strongly Disagree to 5 – Strongly Agree. Item responses were recoded by the HSE team into two composite scores ranging from 0 to 100, with higher scores indicating more positive attitudes.^33^

#### Covariates

Parent-level covariates included: 1) age (less than 44 / 45 or more); 2) cohabitation status (cohabiting with a partner / not cohabiting); 3) education (no qualification or secondary education / A-levels or more, representing any post-secondary education); 4) presence of a mental disorder (yes / no), and; 5) psychological distress (yes / no). Participant-level covariates included: 6) age (13-17 / 18-24); 7) sex (Male / Female); 8) area deprivation (quintiles derived by the HSE team based on the Index of Multiple Deprivation score defined at the Lower Layer Super Output Area level); 9) whether they were in full-time education (yes / no). We did not include participants’ ethnicity and collapsed categories for multiple variables to prevent model convergence issues.

Missingness was low except for GHQ scores and parents’ attitude scores as these were measured in the self-complete questionnaires (see Supplementary Table 4). The comparison of variables across the complete-case (CC) and excluded samples suggested that mothers in the CC sample (n = 630, 78.8% of initial sample) were more likely to have post-secondary education and fathers in the CC sample (n = 428, 76.8% of initial sample) were more likely to have post-secondary education and reside in less deprived areas. There were no statistically significant differences across attitude and mental health variables (see Supplementary Table 5).

### 2.3 Statistical analyses

We first reported the distribution of the main variables in the complete-case samples of participants with data on their mother or father. Using random-intercept Poisson models (nesting participants in households), we then estimated prevalence ratios to assess the associations between having a parent with less prejudice and more tolerance towards mental illness and the two mental health outcomes.^34^ We tested this using a sequential modelling approach, entering: 1) no control variables; 2) the parent’s age, cohabitation status, and education; 3) the parent’s presence of a mental disorder and psychological distress; 4) finally, the participant’s age, sex, area deprivation, and student status. To better interpret results, we plotted the average marginal probabilities from the fully-adjusted models and reported them in the supplementary material.^35^ All descriptive and regression estimates were produced in the complete-case samples, with the weight, strata, and cluster variables provided by the HSE team to account for its sampling design, using Stata 17.^36^

The following sensitivity analyses were produced. First, to explore the potential for bias in using a complete-case approach, we reproduced the main analyses in 20 imputed datasets using Stata’s implementation of multiple imputation by chained equation (using all 13 study variables to build the imputation model).^37^ Second, to account for the fact that multi-level models do not integrate survey weights as standard one-level models, we reproduced the main analyses using scaled versions of the HSE weight using the method A proposed by Carle (2009).^38^ Finally, to examine if parental attitudes had a different magnitude of association across groups, we tested statistical interactions by participants’ age (ages 13-17 vs 18-24) and sex (male vs female). None of the interactions were statistically significant.

## 3. RESULTS

### 3.1. Sample description

Table 1 presents the distribution of sociodemographics in the two samples. The prevalence of a mental disorder among participants was 4.3% and 4.9% in the mother and father samples, respectively. The prevalence of a high level of psychological distress was 13.0% and 10.9% in the mother and father samples. Participants were slightly more likely to be aged 13-17 (56% in the mother sample) and in full-time education (70% in the mother sample).

**TABLE 1.**
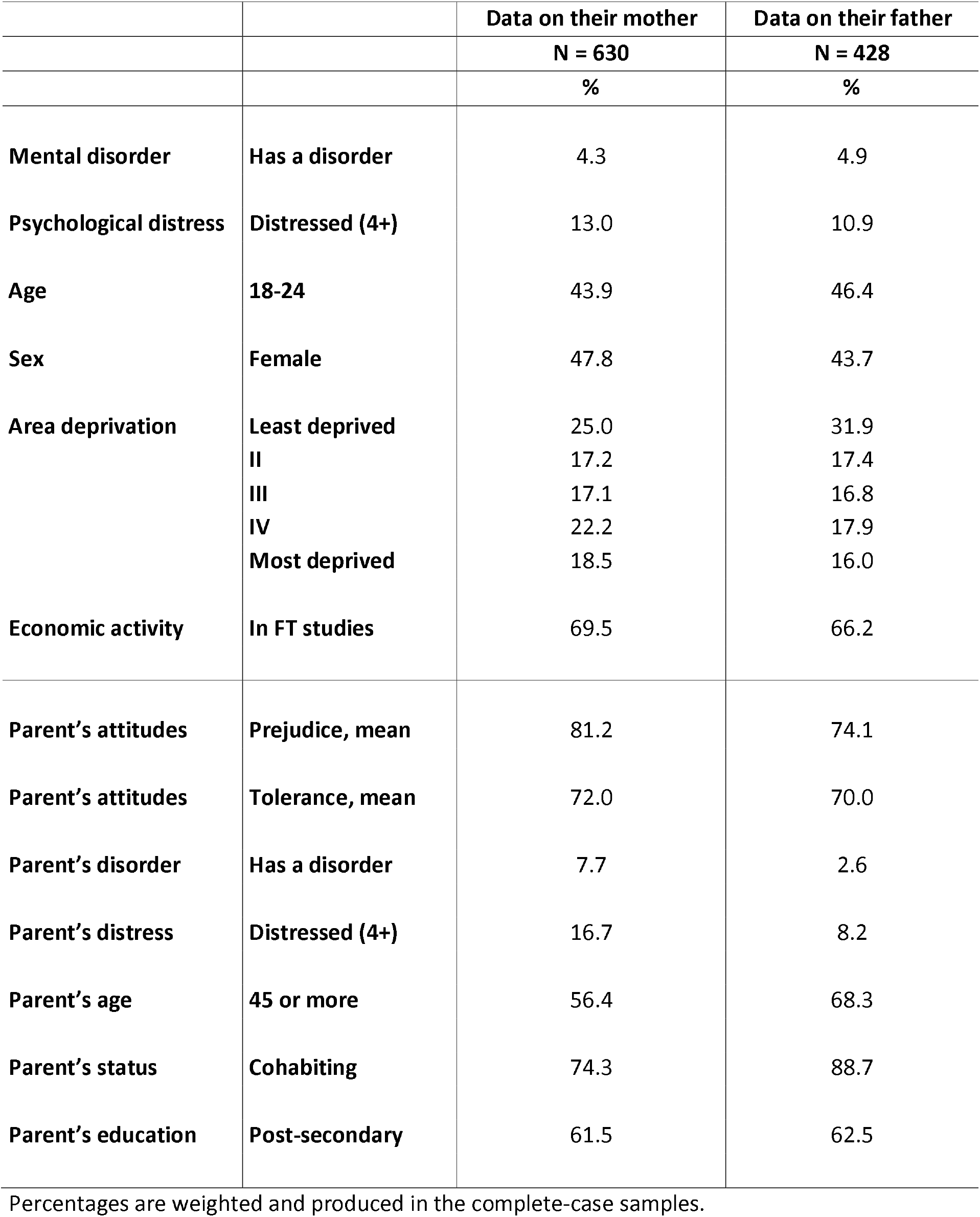
Sample characteristics. Young people aged 13-24 living with parents, Health Survey for England (2014).

|  |  | Data on their mother | Data on their father |
| --- | --- | --- | --- |
|  |  | N = 630 | N = 428 |
|  |  | % | % |
| **Mental disorder** | **Has a disorder** | 4.3 | 4.9 |
| **Psychological distress** | **Distressed (4+)** | 13.0 | 10.9 |
| **Age** | **18-24** | 43.9 | 46.4 |
| **Sex** | **Female** | 47.8 | 43.7 |
| **Area deprivation** | **Least deprived** | 25.0 | 31.9 |
|  | **II** | 17.2 | 17.4 |
|  | **III** | 17.1 | 16.8 |
|  | **IV** | 22.2 | 17.9 |
|  | **Most deprived** | 18.5 | 16.0 |
| **Economic activity** | **In FT studies** | 69.5 | 66.2 |
| **Parent's attitudes** | **Prejudice, mean** | 81.2 | 74.1 |
| **Parent's attitudes** | **Tolerance, mean** | 72.0 | 70.0 |
| **Parent's disorder** | **Has a disorder** | 7.7 | 2.6 |
| **Parent's distress** | **Distressed (4+)** | 16.7 | 8.2 |
| **Parent's age** | **45 or more** | 56.4 | 68.3 |
| **Parent's status** | **Cohabiting** | 74.3 | 88.7 |
| **Parent's education** | **Post-secondary** | 61.5 | 62.5 |

Percentages are weighted and produced in the complete-case samples.

In the mother sample, 4.6% were aged 16-24, 43.3% were aged 25-44, 56.2% were aged 45-64, and 0.1% were aged 65 or more. In the father sample, 5.1% were aged 16-24, 26.6% were aged 25-44, 66.6% were aged 45-54, and 1.6% were aged 65 or more. Mothers had a mean prejudice score of 81.2 (SD = 20.8) and mean tolerance score of 72.0 (SD = 20.4). Fathers had a mean prejudice score of 74.1 (SD = 26.9) and mean tolerance score of 70.0 (SD = 23.5). The correlation between the prejudice and tolerance scores was r = .49 in mothers and r = .45 in fathers. Comparing estimates’ 95% confidence interval across these samples, mothers were more likely to report a more favourable prejudice score (81.2 vs 74.1), mental disorder (7.7% vs 2.6%), and high level of psychological distress (16.7% vs 8.2%), and more likely to be aged less than 45 (43.6% vs 31.7%) and living without a partner (25.7% vs 11.3%) compared to fathers.

### 3.2 Main analyses

#### 3.2.1 Self-reported mental disorder

Table 2 presents the prevalence ratios (PR) for the associations between parents’ prejudice and tolerance scores and their children’s risk of reporting a mental disorder. In the mother sample, both attitude scores were associated with the outcome. In the final model, a one-unit increase was associated with a 3.6% (95%CI 0.7%-6.6%, p = .014) higher relative risk of reporting a mental disorder for the prejudice score and a 3.8% (95%CI 1.1%-6.6%, p = .006) higher relative risk for the tolerance score. In the father sample, only prejudice was associated with the outcome. In the final model, each one-unit increase was associated with a 3.4% (95%CI 0.6%-6.2%, p = .017) higher relative risk of reporting a mental disorder for the prejudice score and a non-significant 2.3% (p = .120) higher relative risk for the tolerance score. Statistical adjustment had a small impact on the magnitude of associations, with the only meaningful change being when controlling for mental health in mothers (prejudice, from PR = 1.047 to PR = 1.037; tolerance, from PR = 1.047 to PR = 1.040).

**TABLE 2.**
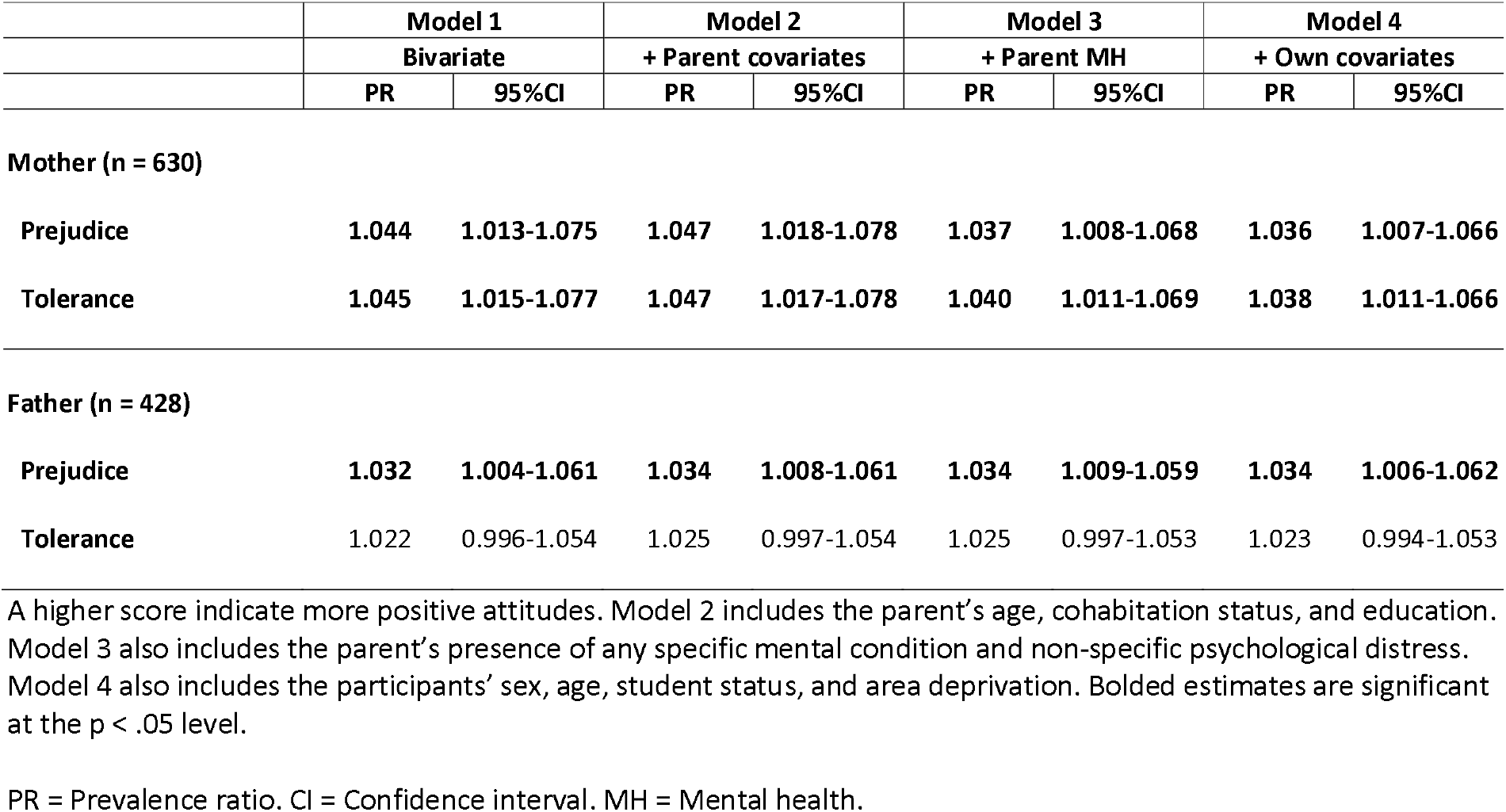
Parents’ attitudes towards mental illness and children’s self-reported presence of a mental disorder. Young people aged 13-24 living with parents, Health Survey for England (2014).

#### 3.2.2 Psychological distress (GHQ-12)

Table 3 presents the prevalence ratios (PR) for the associations between parents’ prejudice and tolerance scores and their children’s risk of reporting a high level of psychological distress. In the mother sample, none of the attitude scores were associated with the outcome. In the final model, a one-unit increase was associated with a non-significant 0.5% (p = .533) higher relative risk of reporting a high level of psychological distress for the prejudice score and a non-significant 0.6% (p = .371) higher relative risk for the tolerance score. In the father sample, only tolerance was associated with the outcome. In the final model, each one-unit increase was associated with a non-significant 0.4% (p = .668) higher relative risk of reporting a high level of psychological distress for the prejudice score and a 2.4% (95%CI 0.8%-4.1%, p = .004) higher relative risk for the tolerance score. Statistical adjustment had no meaningful impact on the magnitude of associations.

**TABLE 3.**
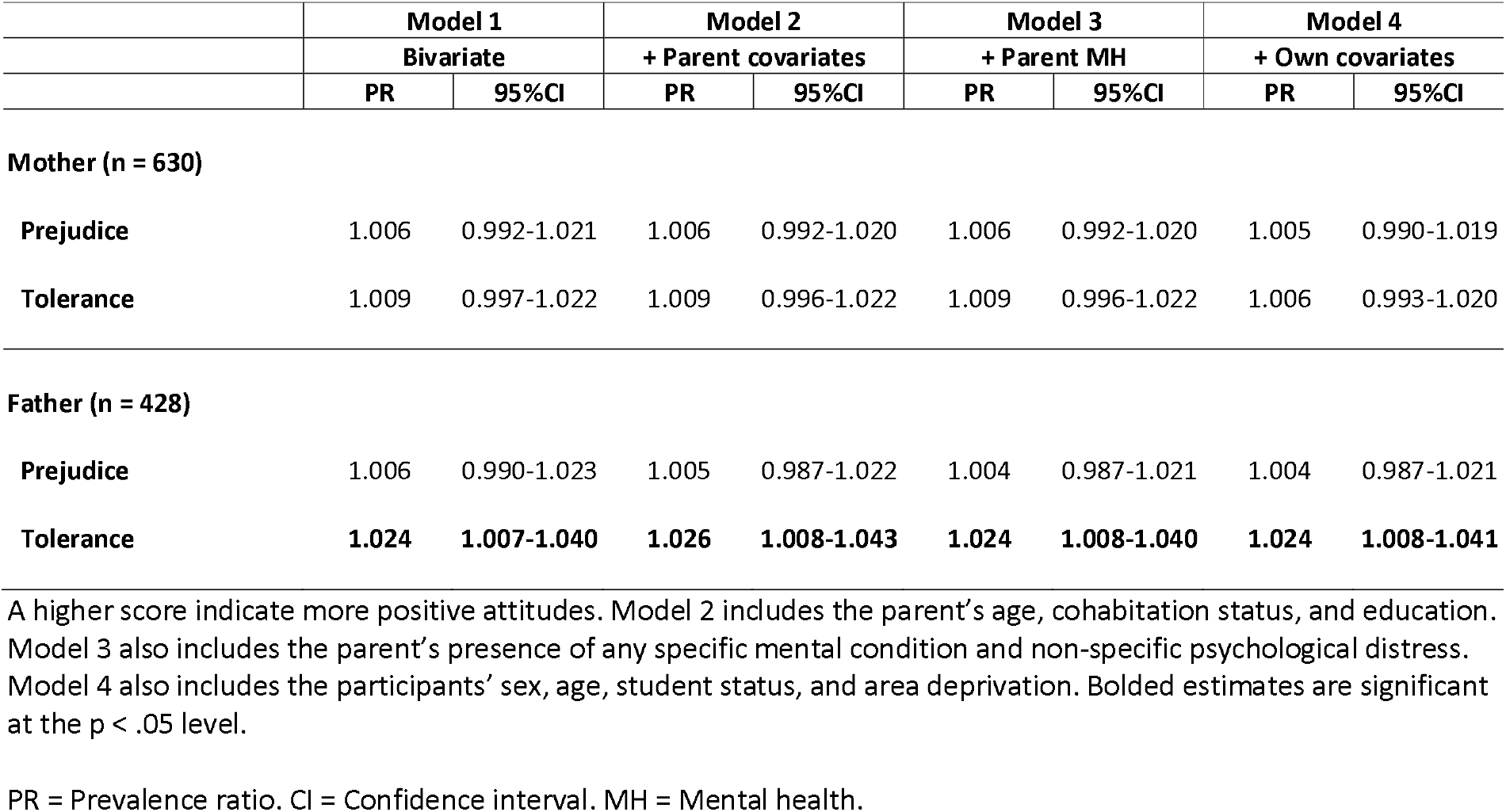
Parents’ attitudes towards mental and children’s self-reported psychological distress. Young people aged 13-24 living with parents, Health Survey for England (2014).

### 3.3 Sensitivity analysis

Analyses were reproduced using multiple imputation and weight scaling to explore the impact of item-level missingness and the integration of standard survey weights in multilevel models, respectively. Multiple imputation yielded consistent results in mothers but weaker associations in fathers that were no longer significant at the .05 level: 1) the fully-adjusted PRs for the association of father’s tolerance score with participants’ risk of self-reporting a specific mental disorder decreased from 1.034 to 1.031 (p = .077); 2) the fully-adjusted PRs for the association of father’s tolerance score with participants’ risk of self-reporting psychological distress decreased from 1.024 to 1.021 (p = .057) (see Supplementary Tables 6 and 7). Using a scaled weight led to slightly weaker effect sizes in both mother and father samples, with no meaningful change in terms of statistical significance (see Supplementary Tables 8 and 9).

## 4. DISCUSSION

In response to the idea that changes in parents’ attitudes may influence the estimation of mental health prevalence among young people over time, we tested the role of mothers’ and fathers’ attitudes towards mental illness in their children’s self-reported mental health in a sample representative of the English population. Overall, the findings support the idea that parents’ attitudes are strongly associated with young people’s self-reported mental health, with differences likely varying in keeping with the parent and mental health indicator used.

Results were very consistent in the mother sample. Mothers who had more favourable attitudes towards mental illness were more likely to have children who self-reported having a specific mental condition. This association was consistent across the two scores of prejudice and tolerance, robust to statistical adjustment (including area deprivation and educational attainment), and only meaningfully affected by considering the mother’s own mental health.^39^ This association, however, did not extend to young people’s non-specific psychological distress. This suggests that the role of the mother’s attitudes may be stronger for specific conditions that are more liable to under-reporting compared with scores derived from a multi-item scale. This may still depend on the framing of the scale within the questionnaire; that is, whether the participant interprets it to be associated with mental illness or not. For reference, the GHQ in the 2014 HSE survey was asked after an impairment screening scale, a quality-of-life scale (which included an item about anxiety and depression), and a general health state continuous scale, and was introduced with the preamble “We would like to know how your health has been in general over the past few weeks”.

Results were less consistent in the father sample, with the main analyses showed that fathers’ prejudice score was associated with the child’s risk of self-reporting a specific condition and tolerance score was associated with the child’s risk of self-reporting non-specific psychological distress. These findings failed to reach statistical significance in the missing data (i.e., imputation) models. Fathers who skipped the mental health items in the self-completion questionnaire may have been more likely to have both less favourable attitudes and children with mental health problems (or vice versa). The weaker consistency of results could also result from the smaller size of the father sample compared to the mother sample, especially given that the magnitude of some associations was similar (e.g, PR = 1.036 in mothers versus PR = 1.034 in fathers for the prejudice score).

The observed association between fathers’ tolerance and their children’ non-specific distress was therefore the more surprising result. It may be that, in parallel to issues related to the self-reporting of a specific mental condition, fathers play a specific role in shaping young people’s awareness about their feelings. Studies focused on emotional socialisation found that, whereas fathers were on average less aware and had less interest in emotions compared with mothers, associations between parents’ emotional-coaching attitudes and child outcomes were actually stronger in fathers.^40,41^ Fully disentangling this, however, would require examining how the father’s role observed here vary as a function of the mother’s attitudes and the sex of the children.^41^

### 4.1. Limitations

Critically, the use of a cross-sectional design precludes us from distinguishing the directionality and causality underlying the associations examined here. Parents’ and children’s health have reciprocal effects over time, including on the parent’s mental health.^42,43^ Studies on the role of poor parental mental health in early life found that the father’s wellbeing was just as important as the mother’s.^44,45^ Studies focused on caregiving for children with mental health problems also found that both parents were impacted by caregiver burden.^46^ Exposure to someone with a mental health problem has also been highlighted as an effective mechanism to change attitudes.^47^ Therefore, it may be the onset of mental health problems in young people that is explaining parents’ more positive attitudes.

The analytic design also tells us little about the life periods in which parents’ attitudes may have a larger impact on their children’s attitudes and behaviours. If this process fully unfolds by the time they become young adults, adjusting for participants’ mental health in adolescence would have substantially attenuated the effect sizes presented here. This issue also affects how we think about the overlap between changes in parents’ attitudes and young people’s mental health over time. That is, the period effect of increasing prevalence estimates of mental health problems that started in the 2010s could capture the cohort effect of growing up in families with less stigmatising attitudes in the 2000s. That said, the evidence does not support improving attitudes before the 2010s in the UK.^48,49^

We note four other limitations. First, our analysis may have missed important confounders of the relationship between parents’ attitudes and young people’s mental health, such as social media use or more detailed data on geography and economic background. Second, the relatively small sample sizes prevented us from reliably testing interactions and the relative contribution of mothers’ and fathers’ attitudes. Third, the linkage of young people with their parents meant that we only used data on those living with parents, which may not capture the reality of those no longer living with them (see differences between those linked with a parent or not in the HSE dataset in Supplementary Table 10). Finally, other relevant outcomes – e.g., whether participants used health care services for a mental health problem – were not measured among participants aged 13-15. Similarly, attitudes were also not measured among participants aged 13-15, preventing us from considering how young people’s attitudes may act as a mediator or effect modifier in the associations investigated here.

### 4.2 Conclusion

The estimation of population levels of mental health among young people may be vulnerable to changes in self-reporting practices over time. These changes may be associated with the improvements in public and interpersonal stigma towards mental illness that have been noted over the past decade.^8,50^ Tackling stigma among parents has been previously identified as a meaningful public health strategy given that many parents are not sufficiently informed regarding mental health.^51^ We found that parents’ attitudes were associated with young people’s mental health, with results being most consistent in mothers and for the self-reporting of specific mental conditions.

This study is part of a larger programme interested in the role of all sources of stigma in young people’s self-reporting practices. At the interpersonal level, other meaningful sources may include peers at school and work, siblings, and other family members. At the structural level, traditional and newer digital media are also likely to play a substantial role.^47^ It is possible that parents act as a channel for these other sources (i.e., mediation), and buffer these sources’ influence depending on their own attitudes (i.e., effect modification). New data collection efforts are needed to assess changes in parents’ attitudes and its role on young people’s self-reported mental health over time.

## Data Availability

All data produced are available online on the UK Data Service.
NatCen Social Research, University College London, Department of Epidemiology and Public Health. (2018). Health Survey for England, 2014. [data collection]. 3rd Edition. UK Data Service. SN: 7919, DOI: 10.5255/UKDA-SN-7919-3

https://www.doi.org/10.5255/UKDA-SN-7919-3

## ACKNOWLEDGEMENTS

The HSE dataset was accessed on the UK Data Service platform. TG holds a Banting Postdoctoral Fellowship award from the Canadian Institutes of Health Research. AM is funded by the UK Economic and Social Research Council (ES/W001454/1).

## ETHICAL STATEMENT

Ethical approval for the 2014 HSE survey was obtained from the Oxford A Research Ethics Committee (reference number 12/SC/0317). No ethical approval was needed for the analysis of the data used here.

