## Supplementary Material for "Associations between parental attitudes towards mental illness and self-reported mental health among young people: Evidence from the Health Survey for England"

**TITLE**

**LAST UPDATED**

October 31^st^, 2022

**Table of contents:**

Figure 1 – Average marginal probabilities for presence of a mental disorder by mother’s attitudes

Figure 2 – Average marginal probabilities for psychological distress by mother’s attitudes

Figure 3 – Average marginal probabilities for presence of a mental disorder by father’s attitudes

Figure 4 – Average marginal probabilities for psychological distress by father’s attitudes

Table 1 – Conditions for “specific mental disorder” in the HSE

Table 2 – GHQ items

Table 3 – CAMI items

Table 4 – Proportion of missingness

Table 5 – Distribution of missingness

Table 6 – Reproducing analyses with multiple imputation: SRMDs

Table 7 – Reproducing analyses with multiple imputation: GHQ

Table 8 – Reproducing analyses with a scaled weight: SRMDs

Table 9 – Reproducing analyses with a scaled weight: GHQ

Table 10 – Differences between youth living with and without parents

**SUPPLEMENTARY FIGURE 1**

**Average marginal probabilities of young people’s presence of a mental disorder by mother’s attitudes towards mental illness. Ages 13-24, HSE 2014 (n = 630).**

|  | Prejudice | Tolerance |
| --- | --- | --- |
| 55 | 1.53474% | 2.02475% |
| 70 | 2.62157% | 3.53518% |
| 85 | 4.47804% | 6.17236% |
| 100 | 7.64918% | 10.7768% |

A higher score indicates a more positive attitude.

Mean prejudice score = 81.2, observed range 20-100.

91.1% of mothers in the CC sample had a prejudice score above 55.

Mean tolerance score = 72.0, observed range 10-100.

84.4% of mothers in the CC sample had a tolerance score above 55.

Average marginal effect for a one-unit increase in the prejudice score = 0.17%, *p* = .013

Average marginal effect for a one-unit increase in the tolerance score = 0.17%, *p* = .004

Models control for parents’ age, education, cohabitation status, presence of a mental disorder, and psychological distres, and participants’ age, sex, area deprivation, and student status.

**N.B.** Differences in a many-unit increase should not be interpreted as in linear regression outputs. Relationships appear non-linear because they come from a non-linear model. This mathematical characteristic is clearest with rare outcomes, as is the case here.

**SUPPLEMENTARY FIGURE 2**

**Average marginal probabilities of young people’s presence of a mental disorder by father’s attitudes towards mental illness. Ages 13-24, HSE 2014 (n = 428).**

|  | Prejudice | Tolerance |
| --- | --- | --- |
| 55 | 2.55813% | 3.70505% |
| 70 | 4.19689% | 5.23259% |
| 85 | 6.88546% | 7.38992% |
| 100 | 11.2964% | 10.4367% |

A higher score indicates a more positive attitude.

Mean prejudice score = 74.1, observed range 0-100.

80.4% of fathers in the CC sample had a prejudice score above 55.

Mean tolerance score = 70.0, observed range 0-100.

77.8% of fathers in the CC sample had a tolerance score above 55.

Average marginal effect for a one-unit increase in the prejudice score = 0.19%, *p* = .035

Average marginal effect for a one-unit increase in the tolerance score = 0.13%, *p* = .156

Models control for parents’ age, education, cohabitation status, presence of a mental disorder, and psychological distres, and participants’ age, sex, area deprivation, and student status.

**N.B.** Differences in a many-unit increase should not be interpreted as in linear regression outputs. Relationships appear non-linear because they come from a non-linear model. This mathematical characteristic is clearest with rare outcomes, as is the case here.

**SUPPLEMENTARY FIGURE 3**

**Average marginal probabilities of young people’s psychological distress by mother’s attitudes towards mental illness. Ages 13-24, HSE 2014 (n = 630).**

|  | Prejudice | Tolerance |
| --- | --- | --- |
| 55 | 11.3476% | 11.4366% |
| 70 | 12.1415% | 12.5588% |
| 85 | 12.9909% | 13.7912% |
| 100 | 13.8997% | 15.1444% |

A higher score indicates a more positive attitude.

Mean prejudice score = 81.2, observed range 20-100.

91.1% of mothers in the CC sample had a prejudice score above 55.

Mean tolerance score = 72.0, observed range 10-100.

84.4% of mothers in the CC sample had a tolerance score above 55.

Average marginal effect for a one-unit increase in the prejudice score = 0.06%, *p* = .535

Average marginal effect for a one-unit increase in the tolerance score = 0.08%, *p* = .372

Models control for parents’ age, education, cohabitation status, presence of a mental disorder, and psychological distres, and participants’ age, sex, area deprivation, and student status.

**N.B.** Differences in a many-unit increase should not be interpreted as in linear regression outputs. Relationships appear non-linear because they come from a non-linear model.

**SUPPLEMENTARY FIGURE 4**

**Average marginal probabilities of young people’s psychological distress by father’s attitudes towards mental illness. Ages 13-24, HSE 2014 (n = 428).**

|  | Prejudice | Tolerance |
| --- | --- | --- |
| 55 | 9.8297% | 6.85624% |
| 70 | 10.3939% | 9.79034% |
| 85 | 10.9904% | 13.9801% |
| 100 | 11.6212% | 19.9628% |

A higher score indicates a more positive attitude.

Mean prejudice score = 74.1, observed range 0-100.

80.4% of fathers in the CC sample had a prejudice score above 55.

Mean tolerance score = 70.0, observed range 0-100.

77.8% of fathers in the CC sample had a tolerance score above 55.

Average marginal effect for a one-unit increase in the prejudice score = 0.04%, *p* = .670

Average marginal effect for a one-unit increase in the tolerance score = 0.25%, *p* = .006

Models control for parents’ age, education, cohabitation status, presence of a mental disorder, and psychological distres, and participants’ age, sex, area deprivation, and student status.

**N.B.** Differences in a many-unit increase should not be interpreted as in linear regression outputs. Relationships appear non-linear because they come from a non-linear model.

**SUPPLEMENTARY TABLE 1**

**Conditions included in the HSE coding frame for self-reported mental disorders**

**04 Mental illness/anxiety/depression/nerves (nes)**

- Alcoholism, recovered not cured alcoholic
- Angelman Syndrome
- Anorexia nervosa
- Anxiety, panic attacks
- Asperger Syndrome
- Autism/Autistic
- Bipolar Affective Disorder
- Catalepsy
- Concussion syndrome
- Depression
- Drug addict
- Dyslexia
- Hyperactive child.
- Nerves (nes)
- Nervous breakdown, neurasthenia, nervous trouble
- Phobias
- Schizophrenia, manic depressive
- Senile dementia, forgetfulness, gets confused
- Speech impediment, stammer
- Stress

**05 Mental handicap**

- Incl. Down's syndrome, Mongol
- Mentally retarded, subnormal

**SUPPLEMENTARY TABLE 2**

**GHQ-12 items**

Have you recently been able to concentrate on whatever you're doing?

Have you recently lost much sleep over worry?

Have you recently felt that you were playing a useful part in things?

Have you recently felt capable of making decisions about things?

Have you recently felt constantly under strain?

Have you recently felt you couldn't overcome your difficulties?

Have you recently been able to enjoy your normal day-to-day activities?

Have you recently been able to face up to problems?

Have you recently been feeling unhappy or depressed?

Have you recently been losing confidence in yourself?

Have you recently been thinking of yourself as a worthless person?

Have you recently been feeling reasonably happy, all things considered?

**SUPPLEMENTARY TABLE 3**

**CAMI-12 items**

*Attitudes statements reflecting views of prejudice and exclusion*

People with mental illness don’t deserve our sympathy

It is frightening to think of people with mental problems living in residential neighbourhoods.

I would not want to live next door to someone who has been mentally ill .

People with mental health problems should not be given any responsibility.

There is something about people with mental illness that makes it easy to tell them from normal people.

One of the main causes of mental illness is a lack of self-discipline and will-power.

*Attitudes statements reflecting views of tolerance and support for community care*

Virtually anyone can become mentally ill.

We need to adopt a far more tolerant attitude toward people with mental illness in our society.

The best therapy for many people with mental illness is to be part of normal community.

Mental illness is an illness like any other.

People with mental health problems are far less of a danger than most people suppose.

Most women who were once patients in a mental hospital can be trusted as baby sitters.

Cronbach’s alpha of mother’s “prejudice” score in the complete-case sample of mothers: 0.74

Cronbach’s alpha of mother’s “tolerance” score in the complete-case sample of mothers: 0.64

Cronbach’s alpha of father’s “prejudice” score in the complete-case sample of fathers: 0.79

Cronbach’s alpha of father’s “tolerance” score in the complete-case sample of fathers: 0.77

**SUPPLEMENTARY TABLE 4**

**PROPORTION of missingness. Ages 13-24, HSE 2014.**

|  | **Data on their mother**  **N = 800** | **Data on their father**  **N = 557** |
| --- | --- | --- |
|  | **%** | **%** |
| **SRMD** | 0 | 0 |
| **GHQ** | 13.4 | 11.5 |
| **Age** | 0 | 0 |
| **Sex** | 0 | 0 |
| **Area deprivation** | 0 | 0 |
| **Economic activity** | 0.4 | 0.2 |
| **Parent’s prejudice** | 10.9 | 17.4 |
| **Parent’s tolerance** | 11.0 | 17.2 |
| **Parent’s SRMD** | 0 | 0 |
| **Parent’s GHQ** | 10.4 | 14.7 |
| **Parent’s age** | 2.0 | 3.6 |
| **Parent’s cohabit.** | 2.0 | 3.6 |
| **Parent’s education** | 2.4 | 4.1 |
| **Listwise deletion** | **21.3** | **23.3** |

Proportions are not weighted.

**SUPPLEMENTARY TABLE 5**

**Distribution of missingness. Ages 13-24, HSE 2014.**

|  |  | **Data on their mother** | | **Data on their father** | |
| --- | --- | --- | --- | --- | --- |
|  |  | **N = 800** | | **N = 557** | |
|  |  | **CC** | **Dropped** | **CC** | **Dropped** |
|  |  | **N = 630** | **N = 170** | **N = 428** | **N = 129** |
|  |  | **%** | **%** | **%** | **%** |
| **SRMD** | **Yes** | 4.3 | 6.4 | 4.9 | 3.9 |
| **GHQ** | **4+** | 13.0 | 11.8 | 10.9 | 8.9 |
| **Age** | **18-24** | 43.9 | 46.3 | 46.4 | 44.3 |
| **Sex** | **Female** | 47.8 | 50.0 | 43.7 | 37.1 |
| **Area deprivation** | **Least deprived** | 25.0 | 15.3 | **31.9** | **19.6** |
|  | **II** | 17.2 | 17.5 | **17.4** | **17.0** |
|  | **III** | 17.1 | 11.4 | **16.8** | **10.6** |
|  | **IV** | 22.2 | 27.1 | **17.9** | **26.9** |
|  | **Most deprived** | 18.5 | 28.7 | **16.0** | **26.1** |
| **Economic activity** | **In FT studies** | 69.5 | 68.3 | 66.3 | 66.2 |
| **Parent’s attitudes** | **Prejudice, mean** | 81.2 | 79.2 | 74.1 | 75.9 |
| **Parent’s attitudes** | **Tolerance, mean** | 72.0 | 75.8 | 70.0 | 60.1 |
| **Parent’s SRMD** | **Yes** | 7.7 | 3.9 | 2.6 | 4.6 |
| **Parent’s GHQ** | **4+** | 16.7 | 7.8 | 8.2 | 10.7 |
| **Parent’s age** | **45 or more** | 56.4 | 53.0 | 68.3 | 66.8 |
| **Parent’s status** | **Cohabiting** | 74.3 | 69.7 | 88.7 | 91.8 |
| **Parent’s education** | **Post-secondary** | **61.5** | **44.8** | **62.5** | **33.1** |

Proportions are weighted. Bolded numbers differ between groups at the *p* < .05 level.

CC = Complete-case.

**SUPPLEMENTARY TABLE 6**

**Testing the role of multiple imputation on associations between parents’ attitudes towards mental illness and children’s self-reported mental health. Ages 13-24, Health Survey for England (2014).**

**Outcome 1: Self-reported mental disorders**

|  | **Model 1** | | **Model 2** | | **Model 3** | | **Model 4** | |
| --- | --- | --- | --- | --- | --- | --- | --- | --- |
|  | **Bivariate** | | **+ Parent covariates** | | **+ Parent MH** | | **+ Own covariates** | |
|  | **PR** | **95%CI** | **PR** | **95%CI** | **PR** | **95%CI** | **PR** | **95%CI** |
|  | **Complete-case** | | | | | | | |
| **Mother (n = 630)** |  |  |  |  |  |  |  |  |
| **Prejudice** | **1.044** | **1.013-1.075** | **1.047** | **1.018-1.078** | **1.037** | **1.008-1.068** | **1.036** | **1.007-1.066** |
| **Tolerance** | **1.045** | **1.015-1.077** | **1.047** | **1.017-1.078** | **1.040** | **1.011-1.069** | **1.038** | **1.011-1.066** |
| **Father (n = 428)** |  |  |  |  |  |  |  |  |
| **Prejudice** | **1.032** | **1.004-1.061** | **1.034** | **1.008-1.061** | **1.034** | **1.009-1.059** | **1.034** | **1.006-1.062** |
| **Tolerance** | 1.022 | 0.996-1.054 | 1.025 | 0.997-1.054 | 1.025 | 0.997-1.053 | 1.023 | 0.994-1.053 |
|  | **Multiple imputation** | | | | | | | |
| **Mother (n = 800)** |  |  |  |  |  |  |  |  |
| **Prejudice** | **1.039** | **1.005-1.075** | **1.044** | **1.011-1.077** | **1.036** | **1.005-1.068** | **1.035** | **1.005-1.067** |
| **Tolerance** | **1.051** | **1.020-1.082** | **1.052** | **1.023-1.083** | **1.046** | **1.017-1.075** | **1.045** | **1.017-1.074** |
| **Father (n = 557)** |  |  |  |  |  |  |  |  |
| **Prejudice** | 1.030 | 0.992-1.070 | 1.034 | 0.998-1.071 | **1.033** | **1.000-1.066** | 1.031 | 0.997-1.067 |
| **Tolerance** | 1.023 | 0.987-1.060 | 1.026 | 0.991-1.063 | 1.025 | 0.991-1.060 | 1.023 | 0.989-1.059 |

Weight was not scaled. Analyses in the MI dataset integrated the weight, psu, and strata variables, but did not use the Stata *subpop()* option.

**SUPPLEMENTARY TABLE 7**

**Testing the role of multiple imputation on associations between parents’ attitudes towards mental illness and children’s self-reported mental health. Ages 13-24, Health Survey for England (2014).**

**Outcome 2: Psychological distress**

|  | **Model 1** | | **Model 2** | | **Model 3** | | **Model 4** | |
| --- | --- | --- | --- | --- | --- | --- | --- | --- |
|  | **Bivariate** | | **+ Parent covariates** | | **+ Parent MH** | | **+ Own covariates** | |
|  | **PR** | **95%CI** | **PR** | **95%CI** | **PR** | **95%CI** | **PR** | **95%CI** |
|  | **Complete-case** | | | | | | | |
| **Mother (n = 630)** |  |  |  |  |  |  |  |  |
| **Prejudice** | 1.006 | 0.992-1.021 | 1.006 | 0.992-1.020 | 1.006 | 0.992-1.020 | 1.005 | 0.990-1.019 |
| **Tolerance** | 1.009 | 0.997-1.022 | 1.009 | 0.996-1.022 | 1.009 | 0.996-1.022 | 1.006 | 0.993-1.020 |
| **Father (n = 428)** |  |  |  |  |  |  |  |  |
| **Prejudice** | 1.006 | 0.990-1.023 | 1.005 | 0.987-1.022 | 1.004 | 0.987-1.021 | 1.004 | 0.987-1.021 |
| **Tolerance** | **1.024** | **1.007-1.040** | **1.026** | **1.008-1.043** | **1.024** | **1.008-1.040** | **1.024** | **1.008-1.041** |
|  | **Multiple imputation** | | | | | | | |
| **Mother (n = 800)** |  |  |  |  |  |  |  |  |
| **Prejudice** | 1.008 | 0.994-1.023 | 1.007 | 0.994-1.021 | 1.007 | 0.993-1.021 | 1.005 | 0.991-1.020 |
| **Tolerance** | 1.013 | 0.999-1.026 | 1.012 | 0.999-1.026 | 1.012 | 0.999-1.025 | 1.010 | 0.996-1.025 |
| **Father (n = 557)** |  |  |  |  |  |  |  |  |
| **Prejudice** | 1.002 | 0.981-1.023 | 1.001 | 0.980-1.022 | 1.001 | 0.982-1.021 | 1.001 | 0.981-1.022 |
| **Tolerance** | 1.020 | 0.999-1.041 | **1.022** | **1.001-1.043** | **1.021** | **1.001-1.041** | 1.020 | 0.999-1.042 |

Weight was not scaled. Analyses in the MI dataset integrated the weight, psu, and strata variables, but did not use the Stata *subpop()* option.

**SUPPLEMENTARY TABLE 8**

**Testing the role of weight scaling on associations between parents’ attitudes towards mental illness and children’s self-reported mental health. Ages 13-24, Health Survey for England (2014).**

**Outcome 1: Self-reported mental disorder**

|  | **Model 1** | | **Model 2** | | **Model 3** | | **Model 4** | |
| --- | --- | --- | --- | --- | --- | --- | --- | --- |
|  | **Bivariate** | | **+ Parent covariates** | | **+ Parent MH** | | **+ Own covariates** | |
|  | **PR** | **95%CI** | **PR** | **95%CI** | **PR** | **95%CI** | **PR** | **95%CI** |
|  | **Weighted, no scaling** | | | | | | | |
| **Mother (n = 630)** |  |  |  |  |  |  |  |  |
| **Prejudice** | **1.044** | **1.013-1.075** | **1.047** | **1.018-1.078** | **1.037** | **1.008-1.068** | **1.036** | **1.007-1.066** |
| **Tolerance** | **1.045** | **1.015-1.077** | **1.047** | **1.017-1.078** | **1.040** | **1.011-1.069** | **1.038** | **1.011-1.066** |
| **Father (n = 428)** |  |  |  |  |  |  |  |  |
| **Prejudice** | **1.032** | **1.004-1.061** | **1.034** | **1.008-1.061** | **1.034** | **1.009-1.059** | **1.034** | **1.006-1.062** |
| **Tolerance** | 1.022 | 0.996-1.054 | 1.025 | 0.997-1.054 | 1.025 | 0.997-1.053 | 1.023 | 0.994-1.053 |
|  | **Weighted, scaled (Method A)** | | | | | | | |
| **Mother (n = 630)** |  |  |  |  |  |  |  |  |
| **Prejudice** | **1.039** | **1.009-1.069** | **1.042** | **1.013-1.072** | **1.034** | **1.005-1.063** | **1.035** | **1.005-1.065** |
| **Tolerance** | **1.041** | **1.013-1.070** | **1.043** | **1.015-1.072** | **1.037** | **1.009-1.065** | **1.036** | **1.009-1.064** |
| **Father (n = 428)** |  |  |  |  |  |  |  |  |
| **Prejudice** | **1.026** | **1.002-1.051** | **1.028** | **1.005-1.052** | **1.028** | **1.005-1.051** | **1.026** | **1.001-1.052** |
| **Tolerance** | 1.018 | 0.994-1.042 | 1.020 | 0.996-1.044 | 1.020 | 0.996-1.044 | 1.018 | 0.993-1.043 |

N complete-case for analysis in mothers = 630 out of 800.

N complete-case for analysis in mothers = 428 out of 557.

**SUPPLEMENTARY TABLE 9**

**Testing the role of weight scaling on associations between parents’ attitudes towards mental illness and children’s self-reported mental health. Ages 13-24, Health Survey for England (2014).**

**Outcome 2: Psychological distress**

|  | **Model 1** | | **Model 2** | | **Model 3** | | **Model 4** | |
| --- | --- | --- | --- | --- | --- | --- | --- | --- |
|  | **Bivariate** | | **+ Parent covariates** | | **+ Parent MH** | | **+ Own covariates** | |
|  | **PR** | **95%CI** | **PR** | **95%CI** | **PR** | **95%CI** | **PR** | **95%CI** |
|  | **Weighted, no scaling** | | | | | | | |
| **Mother (n = 630)** |  |  |  |  |  |  |  |  |
| **Prejudice** | 1.006 | 0.992-1.021 | 1.006 | 0.992-1.020 | 1.006 | 0.992-1.020 | 1.005 | 0.990-1.019 |
| **Tolerance** | 1.009 | 0.997-1.022 | 1.009 | 0.996-1.022 | 1.009 | 0.996-1.022 | 1.006 | 0.993-1.020 |
| **Father (n = 428)** |  |  |  |  |  |  |  |  |
| **Prejudice** | 1.006 | 0.990-1.023 | 1.005 | 0.987-1.022 | 1.004 | 0.987-1.021 | 1.004 | 0.987-1.021 |
| **Tolerance** | **1.024** | **1.007-1.040** | **1.026** | **1.008-1.043** | **1.024** | **1.008-1.040** | **1.024** | **1.008-1.041** |
|  | **Weighted, scaled (Method A)** | | | | | | | |
| **Mother (n = 630)** |  |  |  |  |  |  |  |  |
| **Prejudice** | 1.007 | 0.994-1.021 | 1.006 | 0.993-1.020 | 1.007 | 0.993-1.020 | 1.005 | 0.991-1.019 |
| **Tolerance** | 1.009 | 0.996-1.021 | 1.009 | 0.996-1.022 | 1.009 | 0.996-1.022 | 1.007 | 0.993-1.021 |
| **Father (n = 428)** |  |  |  |  |  |  |  |  |
| **Prejudice** | 1.006 | 0.990-1.022 | 1.005 | 0.988-1.022 | 1.004 | 0.988-1.020 | 1.002 | 0.986-1.019 |
| **Tolerance** | **1.021** | **1.005-1.036** | **1.022** | **1.006-1.039** | **1.021** | **1.005-1.037** | **1.021** | **1.005-1.037** |

N complete-case for analysis in mothers = 630 out of 800.

N complete-case for analysis in mothers = 428 out of 557.

**SUPPLEMENTARY TABLE 10**

**Differences between young people linked with a parent or not. Ages 13-24, HSE 2014.**

|  |  | **Ages 13-24** | | | |
| --- | --- | --- | --- | --- | --- |
|  |  | **N = 1,116** | | | |
|  |  | **Could be linked with mother** | **Could not be linked** | **Could be linked with father** | **Could not be linked** |
|  |  | **N = 800** | **N = 316** | **N = 557** | **N = 559** |
|  |  | **%** | **%** | **%** | **%** |
| **SRMD** | **Yes** | 4.8 | 6.6 | 4.7 | 6.0 |
| **GHQ** | **4+** | 12.9 | 17.1 | **10.6** | **18.0** |
| **Age** | **18-24** | **44.4** | **89.5** | **45.9** | **71.5** |
| **Sex** | **Female** | 48.3 | 49.7 | **42.1** | **55.4** |
| **Area deprivation** | **Least deprived** | 22.9 | 22.6 | **29.1** | **16.4** |
|  | **II** | 17.3 | 11.7 | **17.3** | **13.7** |
|  | **III** | 15.8 | 15.3 | **15.3** | **16.0** |
|  | **IV** | 23.3 | 24.5 | **20.0** | **27.5** |
|  | **Most deprived** | 20.7 | 25.8 | **18.3** | **26.4** |
| **Economic activity** | **In FT studies** | **69.3** | **37.1** | **66.3** | **52.0** |

Proportions are weighted. Bolded numbers differ between groups at the *p* < .05 level.
